# Chronic Disease and Workforce Participation Among Medicaid Enrollees Over 50: The Potential Impact of Medicaid Work Requirements Post-COVID-19

**DOI:** 10.1101/2022.02.07.22270614

**Authors:** Rodlescia S. Sneed, Alexander Stubblefield, Graham Gardner, Tamara Jordan, Briana Mezuk

**Affiliations:** Division of Public Health, Michigan State University, 200 East 1^st^ Street, Flint, MI 48502; Department of Economics, Michigan State University, 486 W. Circle Dr., East Lansing, MI 48824; Department of Epidemiology, University of Michigan School of Public Health, 1415 Washington Heights, Ann Arbor, MI 48109

**Keywords:** Medicaid, Chronic Disease, Health Care Financing/Insurance/Premiums, State Health Policies, Older Adults

## Abstract

As the COVID-19 pandemic wanes, states may reintroduce Medicaid work requirements to reduce enrollment. Using the Health and Retirement Study, we evaluated chronic disease burden among beneficiaries aged >50 (n=1460) who might be impacted by work requirements (i.e. working <20 hours per week). Seven of eight chronic conditions evaluated were associated with reduced workforce participation, including history of stroke (OR: 7.35; 95% CI: 2.98-18.14) and lung disease (OR: 4.39; 95% CI: 2.97-7.47). Those with more severe disease were also more likely to work fewer hours. Medicaid work requirements would likely have great impact on older beneficiaries with significant disease burden.

**Key Points:**

- Chronic disease linked to reduced work among older Medicaid beneficiaries.
- Work requirements would greatly impact those aged >50 with chronic conditions.
- Coverage loss would have negative implications for long-term disease management.

## INTRODUCTION

Medicaid is the primary public health insurance program for low-income Americans. It plays a central role in our health care system, providing health insurance for one in five Americans (Kaiser Family Foundation, 2019), most of whom lack access to other affordable health insurance options. Additionally, it provides more than 50% of long-term care financing, and helps to pay for premiums and cost-sharing for 10 million low-income Medicare beneficiaries (Rudowitz et al., 2019).

Given its critical role in our nation’s health insurance infrastructure, Medicaid also accounts for a significant portion of federal and state expenditures. In FY 2020, total Medicaid program expenditures exceeded $652 billion, up from $597 billion in FY 2019 (Medicaid.gov, n.d.). This spending growth can be largely attributed to the COVID-19 pandemic, which has led to a significant increase in Medicaid enrollment and subsequent spending. All 50 states have reported pandemic-related fiscal stress, as demands for health insurance have increased while state revenues have declined (Hinton et al., 2021). From February 2020 to June 2021, Medicaid enrollment increased 16.8%, which corresponds to 12 million new enrollees (Corallo, 2022). Consequently, states have found themselves facing difficult budget decisions to meet balanced budget requirements (Gifford et al., 2020).

Historically, fiscal stress to the Medicaid system has resulted in policy efforts to reduce spending. For example, the Deficit Reduction Act of 2005 gave states authority to require proof of citizenship for Medicaid beneficiaries, charge premiums for children and families between 100 and 150% of the Federal Poverty Level (FPL), and introduce cost sharing for families with income more than 150% of the FPL (Markus & Rosenbaum, 2006). States have also used eligibility and enrollment restrictions as a strategy for reducing costs. For example, in October 2020, the Centers for Medicare and Medicaid Services (CMS) approved the state of Indiana’s request to disenroll and lock-out beneficiaries for non-payment of premiums and to lock-out beneficiaries who did not renew their eligibility in a timely fashion (Kaiser Family Foundation, 2022). Additionally, at least 8 states have applied for waivers to Medicaid’s retroactive coverage requirements, which allow people to receive coverage for health services received in the three months prior to filing a Medicaid application if they met the eligibility requirements at the time (Cuello, 2021; Shafer et al., 2020).

One recent attempt to curb Medicaid spending has been the introduction of work requirements. From 2018-2020, nineteen states (with guidance from the Trump administration and CMS) drafted policies to impose work requirements among working-aged Medicaid beneficiaries as a condition of enrollment. Proponents argued that these requirements promoted financial independence for families and individuals by increasing employment, shifting Medicaid recipients to employer-based health care coverage and reducing Medicaid costs (Medicaid and CHIP Payment and Access Commission, 2017). Opponents argued that work requirement policies were largely untested and that they could disenfranchise the nation’s most medically-vulnerable citizens (Garfield et al., 2019a). Although Medicaid work requirements were originally sanctioned by CMS under the Trump administration, they were later rescinded in 2021 with the incoming Biden administration.

While no states are currently implementing work requirements, as states attempt to recover economically from the COVID-19 pandemic, strategies to reduce Medicaid enrollment are likely to reemerge. Further, The COVID-19 pandemic has resulted in a widespread shift in the U.S. labor force, such that there is an ongoing trend of employees voluntarily leaving their jobs at record numbers (i.e. the “Great Resignation”; Washington Post Live; 2021). With this, Medicaid work requirements may re-emerge as a policy conversation in an attempt to both reduce Medicaid enrollment and to rebuild the U.S. post-pandemic workforce. Thus, it is still important to consider the potential impact of Medicaid work requirement policies in policy analyses.

We were specifically interested in studying the potential impact of Medicaid work requirements on adults aged >50. Medicaid beneficiaries in this subgroup are of particular interest, as they likely have significant age-related chronic disease burden. More than 50% of individuals in the general population of adults ages 51-64 have two or more chronic health conditions (Buttorff et al., 2017), and rates of chronic disease have increased significantly in the last quarter century among adults in this group (United Health Foundation, 2016). Burden of disease may be even higher among Medicaid recipients, as management of chronic health conditions requires ongoing partnership with a primary care provider, which is greatly impacted by access to adequate health care coverage (Garfield et al., 2019b). We know little about the specific health needs of Medicaid enrollees who are not in the workforce. Additionally, we know little about the true impact of chronic disease in this population with respect to both disease severity and healthcare utilization. Thus, our goal was to evaluate chronic disease burden among Medicaid beneficiaries ages 51-64 who might be subject to Medicaid work requirements. Additionally, we were interested in differences in disease severity and healthcare utilization between Medicaid beneficiaries with reduced workforce participation and those working at higher levels.

Our population of interest included Medicaid recipients ages 51-64 working less than 20 hours per week (the cutoff used by most states in their Medicaid demonstration projects). We addressed our research questions using data from the Health and Retirement Study, a nationally-representative study of U.S. community-dwelling adults aged >50. We compared individuals working less than 20 hours per week to their counterparts working at least 20 hours per week.

## METHODS

### Participants and Study Design

We used data from the 2016 wave of the Health and Retirement Study (HRS), a biennial, longitudinal panel study of community-dwelling adults aged >50 (University of Michigan, 2016; University of Michigan, 2018). The survey uses a nationally-representative, multi-stage area probability survey design of U.S. households, oversampling Blacks, Hispanics, and Florida residents (Juster & Suzman, 1995). This current study was exempt from Michigan State University IRB review.

The 2016 HRS wave included 20,912 individual participants. From this sample, we excluded participants who were not ages 51-64 (n=11,053) and those not on Medicaid (n=8,107). We also excluded 202 participants who receiving Supplemental Security Income (SSI), as these individuals would likely be excluded from Medicaid work requirements due to disability. Additionally, we excluded those with missing data on workforce participation (n=71), and those missing data on one of our standard covariates (age, race/ethnicity, marital status, gender, education; n=19). Our final sample included 1,460 participants who were 46.53% Non-Hispanic White, 21.78% Hispanic, 21.33% Non-Hispanic Black, and 10.36% other racial/ethnic backgrounds. Participants were ages 51-64 (mean age 56.79, SD4.22) and 58.56% female (Table 1).

**Table 1.** Demographic Characteristics and Health Status of Study Sample

|  | Total Sample | Working < 20 hours per week | Working ≥ 20 hours per week | p-value |
| --- | --- | --- | --- | --- |
| Variable | % | % | % |  |
| Mean Age (SD<sup>a</sup>) | 56.79 (4.22) | 57.06 (4.30) | 56.03 (3.89) | 0.01 |
| Race |  |  |  | 0.23 |
| Non-Hispanic White | 46.53 | 45.21 | 50.25 |  |
| Non-Hispanic Black | 21.33 | 23.43 | 15.46 |  |
| Hispanic | 21.78 | 21.68 | 22.07 |  |
| Non-Hispanic Other | 10.36 | 9.69 | 12.22 |  |
| % Female | 54.80 | 56.61 | 49.71 | 0.18 |
| Education |  |  |  | 0.02 |
| Less than High School | 33.35 | 28.32 | 17.84 |  |
| High School Degree | 25.57 | 31.57 | 38.34 |  |
| GED<sup>b</sup> | 10.03 | 11.19 | 6.78 |  |
| Some College | 13.38 | 13.55 | 12.90 |  |
| College Degree or Higher | 17.67 | 15.37 | 24.14 |  |
| Marital Status |  |  |  | 0.15 |
| Married | 36.25 | 34.94 | 39.92 |  |
| Partnered | 8.62 | 9.38 | 6.48 |  |
| Separated/Divorced | 30.16 | 28.38 | 35.15 |  |
| Widowed | 6.13 | 6.58 | 4.86 |  |
| Never Married | 18.84 | 20.72 | 13.59 |  |
| Mean number of chronic health conditions (SD) | 2.55 (1.90) | 2.91 (1.89) | 1.53 (1.44) | <.0001 |
| Number of chronic health conditions |  |  |  | <.0001 |
| 0 conditions | 12.03 | 7.17 | 25.65 |  |
| 1 Condition | 18.23 | 14.40 | 28.95 |  |
| 2 Conditions | 21.40 | 20.24 | 24.64 |  |
| 3 or more conditions | 48.35 | 58.19 | 20.76 |  |

<sup>a</sup>Standard Deviation; <sup>b</sup>General Educational Development

### Assessment of Workforce Participation

Workforce participation was evaluated based on self-report. Participants were asked about the number of hours they worked per week in a primary or secondary job. Participants were considered to have reduced workforce participation if they worked less than 20 hours per week across jobs.

### Assessment of Chronic Health Conditions

We evaluated the prevalence of the following eight self-reported chronic conditions: diabetes, hypertension, cancer, lung disease, heart disease, stroke, psychiatric problems, and arthritis. These conditions were each assessed as a binary (yes/no) indicator and were summed to create an indicator of cumulative disease burden (range: 0 to 8). Analyses were conducted using the number of chronic health conditions as a continuous variable as well as a categorical variable (categories: 0 conditions, 1 condition, 2 conditions, 3 or more conditions).

### Indicators of Disease Severity

The severity of four of these conditions (arthritis, diabetes, hypertension, and lung disease) was also assessed by self-report via the following questions.

#### Arthritis severity

To assess arthritis severity, participants were asked about use of opioid pain medication in the last three months (yes/no), use of over the counter pain medication in the past 3 months (yes/no), trouble with pain (yes/no), if their arthritis had gotten better worse or stayed the same in the last 2 years, and if they had surgery or joint replacement because of arthritis in the past 2 years (yes/no)

#### Diabetes severity

Participants were asked to rate their eyesight using glasses or corrective lenses (excellent, very good, good, fair, or poor), if they were currently using insulin shots or a pump (yes/no), if they had persistent swelling in the feet or ankles since the last interview (yes/no), and if they had stayed overnight in the hospital in the past two years (yes/no).

#### Hypertension severity

Participants were asked if they were currently taking blood pressure medications (yes/no), if they had stayed overnight in the hospital in the past two years (yes/no), and if they had ever had a stroke (yes/no).

#### Lung Disease Severity

Participants were asked if they were currently taking medications or receiving treatment for a lung condition (yes/no), if they were receiving oxygen for the lung condition (yes/no), if the lung condition had changed since the last interview (better, worse, stayed the same), and if they had been hospitalized over the last two years (yes/no).

### Covariates

Demographic variables included in all analyses were age (continuous variable), sex (male/female), education (no degree, high school diploma, GED [general educational development], two-year degree/some college, four-year degree, master’s degree, professional degree), race (Non-Hispanic White, Non-Hispanic Black, Hispanic, Non-Hispanic Other), and marital status (Married, Partnered & Unmarried, Separated, Divorced, Widowed, Never Married).

### Statistical Analyses

Given the complex sample design of the HRS (Ofstedal et al., 2011), all analyses were conducted using the survey procedures in Stata 16 (Stata, n.d.a). We evaluated the association between chronic health conditions and reduced workforce participation, defined as working less than 20 hours per week. We used logistic regression to determine the association between each of the eight chronic health conditions and reduced workforce participation, calculating odds ratios (OR) and 95% confidence intervals (CIs). All analyses adjusted for demographic characteristics.

We also evaluated the association between cumulative burden of health conditions (possible range 0 to 8) and reduced workforce participation. All analyses adjust for our standard control variables.

To evaluate differences in indicators of disease severity based on workforce participation, we performed separate analyses among all individuals with the four conditions that assessed severity: diabetes, high blood pressure, arthritis, and lung disease. We used the margins command in Stata (Stata, n.d.b) to calculate probabilities for each indicator based on hours worked per week (<20 hours versus 20+ hours), adjusting for age, sex, race/ethnicity, education, and marital status.

## RESULTS

Of the 1460 Medicaid recipients aged 51 to 64 in our analyses, 66.31% worked less than 20 hours per week and 33.69% worked at least 20 hours per week (Table 1). Of those working less than 20 hours per week, 93.89% did not work at all. Those who were employed in this group worked a mean of 11.9 hours per week (SD 5.3). About 26.3% of the sample worked at least 20 hours per week. In this group, the mean number of hours worked per week was 39.7 hours per week (SD 15.8). Demographic predictors of reduced workforce participation included older age, less than a high school education, having a GED, and being partnered but unmarried. Notably, neither gender nor race/ethnicity were associated with reduced workforce participation.

Chronic disease burden was high in the sample. Overall, about 32% of the sample had diabetes, 10% had cancer, 21% had lung disease, 23% had heart problems, 12% have had a stroke, 39% had a psychiatric condition, 55% had arthritis, and 62% had hypertension. Only 12.0% had none of the eight health conditions and nearly half (48.35%) had three or more of these conditions. Seven of the eight chronic health conditions (hypertension, diabetes, heart disease, lung disease, stroke, psychiatric problems, and arthritis) were individually associated with increased odds of reduced workforce participation (Figure 1). The strongest associations with reduced workforce participation were observed for history of stroke (OR: 7.35; 95% CI: 2.98-18.14) and lung disease (OR: 4.39; 95% CI: 2.97-7.47). A history of cancer was not associated with reduced workforce participation (OR: 1.43; 95% CI: 0.65-3.15). Individuals with at least two chronic health conditions had greater odds of reduced workforce participation than those with less than two chronic conditions (OR: 4.02; 95% CI: 2.68-6.02). Those with three or more conditions had greater odds of reduced workforce participation than those with less than three chronic conditions (OR: 5.30; 95% CI 3.17-8.87).

**Figure 1.**
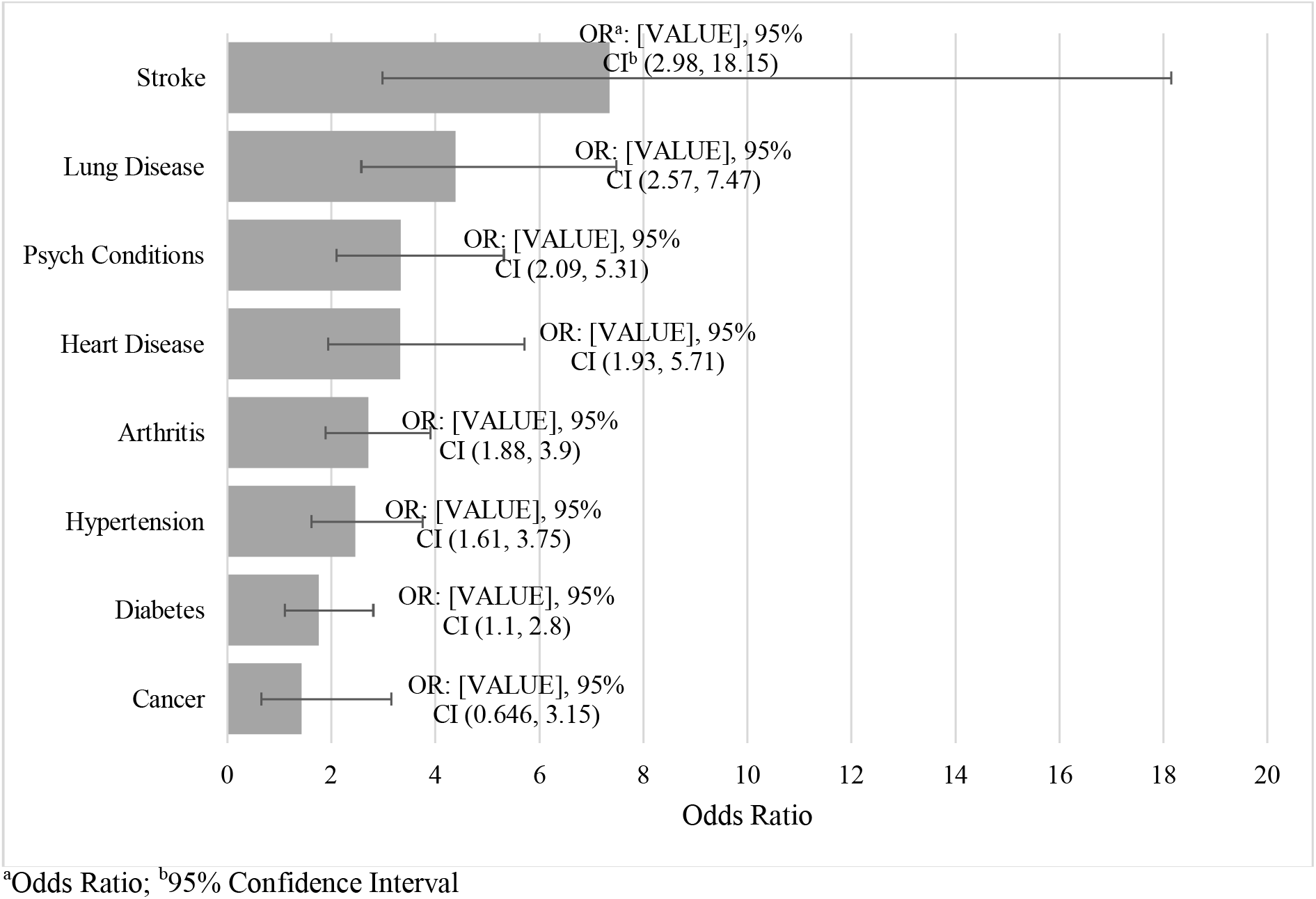
Association between prevalent chronic health conditions and reduced workforce participation (working <20 hours per week) among Medicaid Beneficiaries ages 51-64. Each chronic health condition was evaluated in a separate logistic regression model, adjusting for age, race/ethnicity, sex, marital status, and education.

### Indicators of Disease Severity and Reduced Workforce Participation

We examined the relationship between disease severity and workforce participation for four health conditions that assessed these indicators. Among those with diabetes, 36.4% of those who worked < 20 hours per week reported insulin use, compared to only 22.2% of those who worked ≥ 20 hours per week (p=0.04; Figure 2). Additionally, 46% of those with diabetes working < 20 hours per week reported swelling in their feet or ankles compared to only 22% of those who worked ≥ 20 hours per week (p=.03). There were no significant differences in rates of hospitalization or prevalence of fair/poor eyesight based on workforce participation. Among those with arthritis, fifty percent of those working <20 hours per week reported worsening of their arthritis in the past 2 years, compared to only 26% of those working 20+ hours (p=0.03; Figure 3). Additionally, those working <20 hours were more likely than their counterparts to report difficulty climbing a flight of stairs (48% versus 23%; p=0.001), difficulty walking one block (42% versus 14%; p<0.0001), and use of opioids (41% versus 24% p=0.01). There was no association between reduced workforce participation and self-reported trouble with pain or use of over-the-counter pain medications among those with arthritis.

**Figure 2.**
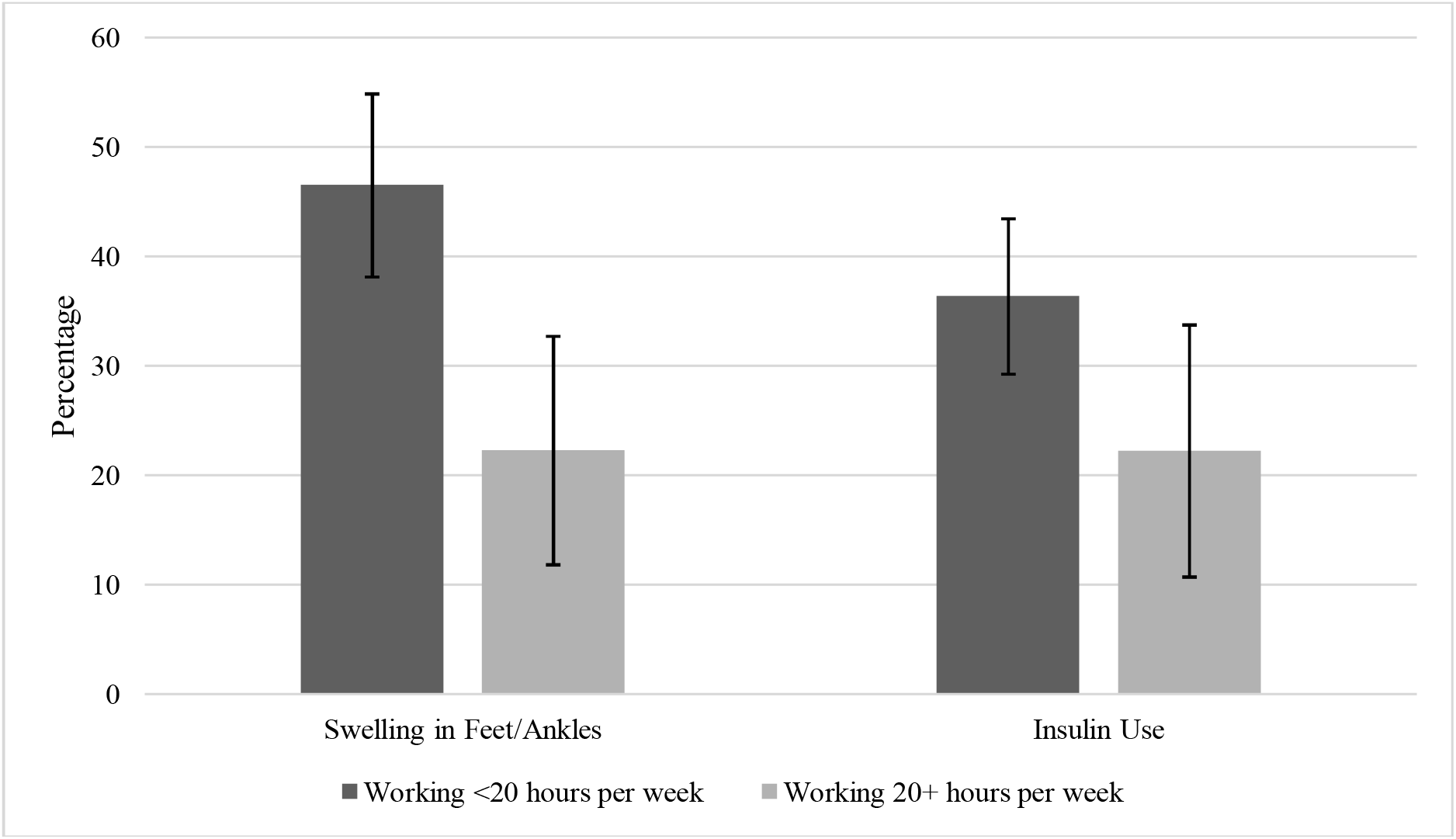
Self-Reported Indicators of Disease Severity and Workforce Participation Among Those with Diabetes Error bars represent the 95% confidence intervals. Analyses adjusted for age, race/ethnicity, sex, marital status, and education.

**Figure 3.**
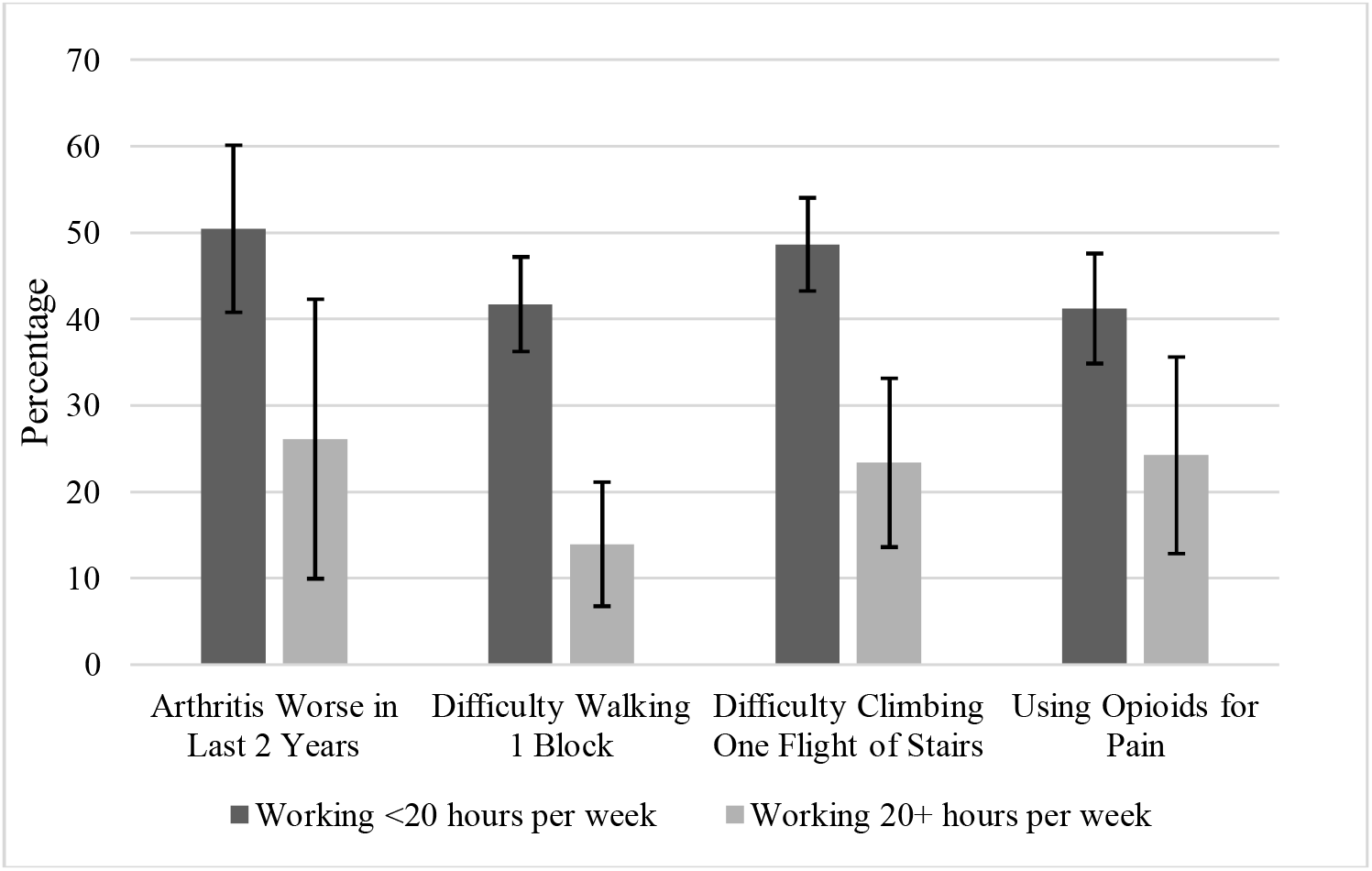
Self-Reported Indicators of Disease Severity and Workforce Participation Among Those with Arthritis Error bars represent the 95% confidence intervals. Analyses adjusted for age, race/ethnicity, sex, marital status, and education.

**Figure 4.**
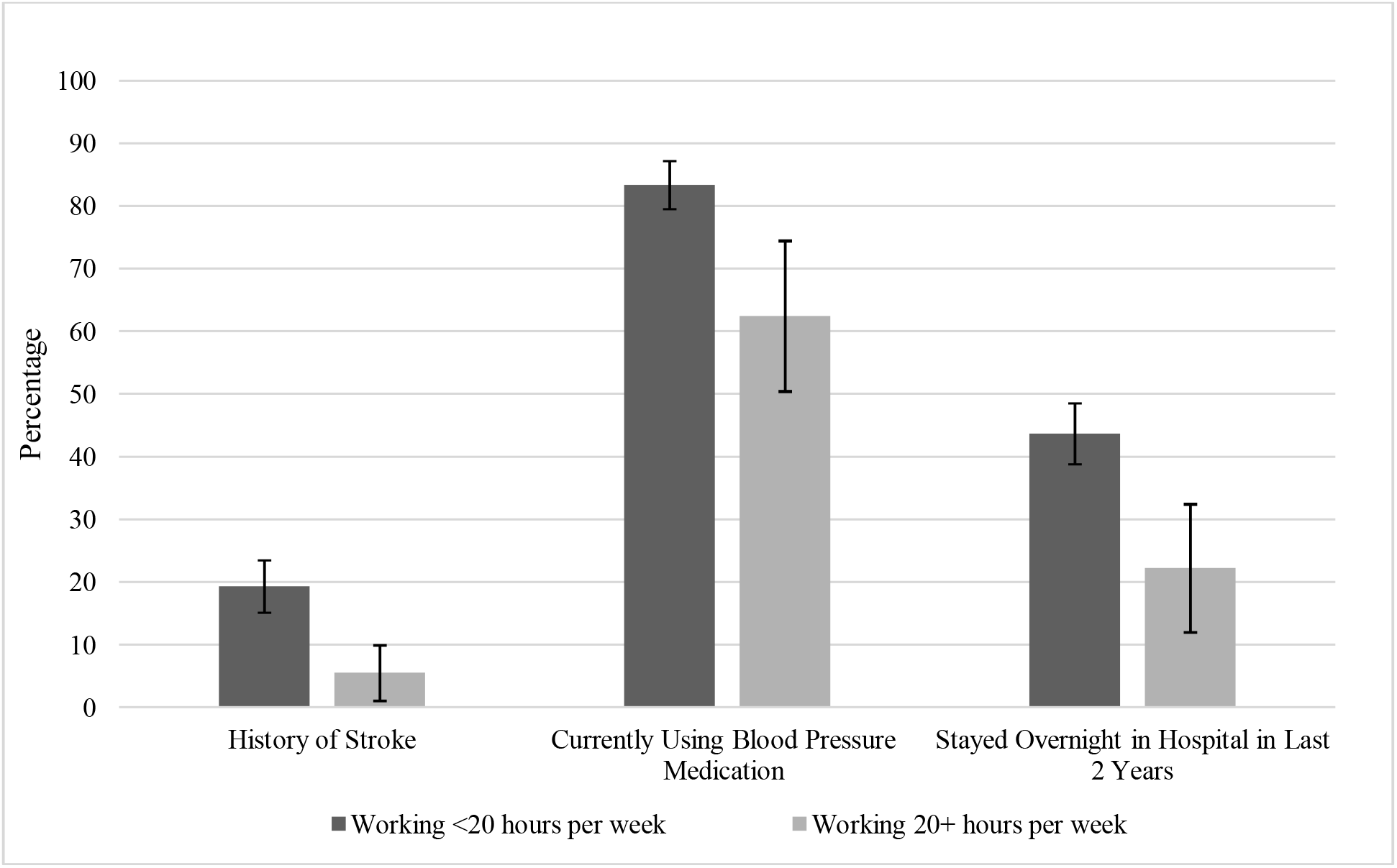
Self-Reported Indicators of Disease Severity and Workforce Participation Among Those with Hypertension Error bars represent the 95% confidence intervals. Analyses adjusted for age, race/ethnicity, sex, marital status, and education.

Among those with a lung condition, 73% of those working <20 hours per week were currently using medication or treatment for the condition, compared to only 43% of those working 20+ hours per week (p=0.008). There was no statistically significant difference among those working <20 hours versus those working 20+ hours with respect to hospitalization in the last 2 years (41% versus 23%; p=0.16), oxygen use (15% versus 4%; p=0.19) or worsening of the lung condition in the last 2 years (26% versus 20%; p=0.69).

Finally, among those with hypertension, 83% of those working <20 hours per week were currently taking blood pressure medications, compared to only 62% of those working 20+ hours per week. Additionally, 43% of those working <20 hours per week had been hospitalized in the last 2 years, compared to only 22% of those working 20+ hours per week (p=0.002). Further, 19% of those working <20 hours per week reported a history of stroke, compared to only 5% of those working 20+ hours per week.

## DISCUSSION

We observed that chronic disease burden is strongly associated with reduced workforce participation among Medicaid beneficiaries aged 51 to 64. Individuals with reduced workforce participation had greater odds of disease for seven of eight chronic conditions, even after controlling for variables commonly associated with workforce participation, including age, race/ethnicity, sex, marital status, and education. We also observed that, among those with individual chronic health conditions, those with reduced workforce participation had poorer condition-related outcomes, suggesting greater disease severity. Overall, our findings suggest that work requirements for Medicaid beneficiaries in this age group would have the greatest impact on those with high disease burden. This has broad implications for population health, as individuals who have multiple chronic health conditions report greater healthcare utilization than their counterparts in the form of more doctor’s visits, prescriptions, and referrals to specialists (Buttorff et al., 2017). Loss of coverage for these important services would likely have negative implications for long-term disease management.

Our findings are consistent with other studies in this area. The Kaiser Family Foundation reported that Medicaid beneficiaries ages 19-64 who reported poor health were 50% less likely to be employed as those reporting excellent or good perceived health, rendering them more likely to face coverage losses (Garfield et al., 2018a). Additionally, the Kaiser Family Foundation reported that 35% of those not working have multiple chronic conditions, including high blood pressure, high cholesterol, arthritis, or heart disease (Garfield et al., 2018b). Notably, despite their health problems, these beneficiaries were not currently receiving social security income, which is typically associated with disability. Further, data from the Michigan Medicaid expansion program demonstrated that 74% of Medicaid beneficiaries ages 19-64 who were out of work had at least one chronic health condition, compared to only 62% of those who were currently employed (Tipirneni et al., 2018).

Our findings are consistent with a broader literature linking poor health to less employment. Numerous studies have demonstrated that poor health is associated with greater risk of exit from paid employment (van Rijn et al., 2013). Further, among those who remain in the workforce, chronic disease has been strongly linked to limitations in individuals’ abilities to perform both the physical and psychosocial demands of work (Lerner et al., 2000). People with chronic health conditions report less productivity and more difficulty in performing physical work-related tasks (Jinnett et al., 2017). Additionally, individuals with chronic conditions face more workplace discrimination and less employer support than their counterparts (Siu et al., 2012).

### Limitations

This study is not without limitations. We utilized data from the 2016 HRS to evaluate associations between chronic disease and reduced workforce participation. These data were collected prior to the COVID-19 pandemic, which has had known impacts on both employment and Medicaid enrollment (Hinton et al., 2021). Consequently, our sample may not reflect the current population of adults aged >50 receiving Medicaid benefits. Despite this, our study still has a number of strengths. We utilized data from a population-based sample including more than 20,000 individual participants. This provided us with an adequate subsample of Medicaid beneficiaries over 50 to perform our analyses. Further, the Health and Retirement study includes a large battery of health-related information, which allowed us to characterize the health status of our sample in great detail.

## CONCLUSION

This work has important implications for Medicaid policy. Although Medicaid work requirements are not currently being implemented anywhere in the U.S., conversations related to this issue may re-emerge as states attempt to recover from the financial impact of the pandemic. In December 2021, CMS contacted the states with approved work requirements through drafted letters, saying workforce requirements among Medicaid beneficiaries did not promote Medicaid objectives (Kaiser Family Foundation, 2022). Six states, however, still have approved applications for implementing work requirements that have not yet been acted on by CMS (Kaiser Family Foundation, 2022), and there is still great debate over whether Medicaid benefits should be tied to employment. As CMS and the states consider next steps on work requirements, our findings suggest that any decisions would have great impact on Medicaid beneficiaries over 50 who have chronic health conditions.

## Data Availability

All data are available online at: https://hrsdata.isr.umich.edu/data-products/rand-hrs-longitudinal-file-2018 and https://hrsdata.isr.umich.edu/data-products/2016-hrs-core

## Acknowledgments

This work was supported by the Robert Wood Johnson Foundation’s (RWJF) Policies for Action program under grant number 77342. This is a secondary analysis that uses data from the Health and Retirement Study, (2016 HRS Core and RAND HRS Longitudinal File 2018), sponsored by the National Institute on Aging under grant number NIA U01AG009740 and conducted by the University of Michigan.

